# Covid-19 and Mental Health of Individuals with Different Personalities

**DOI:** 10.1101/2021.05.24.21257581

**Authors:** Eugenio Proto, Anwen Zhang

## Abstract

Several studies have been devoted to establishing the effects of the Covid-19 pandemic on mental health across gender, age and ethnicity. However, much less attention has been paid to the differential effect of lockdown according to different personalities. We do this using the UKHLS longitudinal dataset, representative of the UK population. The UKHLS dataset allows us to assess the mental health of the same respondent during the Covid-19 period and the year before based on their personality “Big Five” traits and cognitive skills. We find that during the Covid-19 period individuals who have more Extrovert and Open personality report a higher mental health deterioration, while the ones scoring higher in Agreeableness are less affected. The effect of Openness is particularly strong: one more standard deviation predict one more symptom on the GHQ12 test for about 1 respondent over 4. In female respondents, Cognitive Skills and Openness are particularly strong predictors of deterioration. Neuroticism seems to predict more mental health deterioration, as it is normal to expect, but this effect is not significant in the main specifications of the estimated model. The study’s results are robust to the inclusion of potential confounding variables such as changes in: physical health, household income and job status (like unemployed or furloughed).

## 1 Introduction

The question of whether Covid-19 affects the mental health of different individuals in differently ways is very open and compelling. Several studies have been devoted to establishing the effects on different age, gender and ethnicity (e.g. Banks and Xu, 2020; Daly et al., 2020; Davillas and Jones, 2020; Etheridge and Spantig, 2020; Proto and Quintana-Domeque, 2021). However, little or no attention has been paid to the differential effect of Covid-19 according the differences in individual personalities.

Analyzing the differential effect of the pandemic according to personality is important at least for three reasons. First, it can lead to identification of at-risk groups as well as more personalized psychological or psychiatric treatments, even for the post-Covid period. Second, understanding how individuals with different personality react to an extreme condition like a lockdown can shed more light on the link between personality and mental health. Third, it can make clearer unintended consequences of Covid-19 restrictions and inform policy-making.

The Covid-19 period can be thought as a natural experiment where a sort of stress test is naturally induced. The UKHLS provides longitudinal data for the same sample of individuals representative of UK population from before and during the Covid-19 period, where individual mental health is monitored before and during the event. Furthermore, the UKHLS dataset provides the necessary information about personality traits and cognitive skills that are the main explanatory variables in the current study. Hence the UKHLS is an ideal tool to analyze effects of this pandemic on mental health deterioration among individuals with different personalities.

Some confounding factors are potentially relevant in our study. We show that our results are robust to the inclusion of controls such as changes in: physical health, household income, job status (like unemployed or furloughed), marital status, household size and geographic location, during the Covid-19 period.

There is a widespread consensus on the personality classification based on the OCEAN five-factor model, or Big Five Goldberg (1993); Digman (1994); Markon et al. (2005); Costa Jr and McCrae (2008). And, following this classification, there is a large literature analyzing the link between personality and mental health (e.g. Watson and Clark, 1994; Krueger and Tackett, 2003; Clark, 2005).^1^ Further, there are several contributions studying how personality affects self-reported subjective wellbeing (e.g. Watson and Clark, 1992; Diener and Lucas, 1999; Boyce and Wood, 2011; Proto and Rustichini, 2015). We show that the data used in the current study produce results that are consistent with these contributions. Building on this literature, to the best of our knowledge, we are the first to show with real non-experimental data how an external shock interacts with personality to affect mental health.

We find that during the Covid-19 period individuals who have more Extrovert and Open personality report a higher mental health deterioration, while the ones scoring high in Agreeableness are less affected. The effect of Openness is particularly strong and seems increasing in magnitude thorough the entire period.

Neuroticism seems to predict more mental health deterioration, but this effect is not significant in the main specifications of the estimated model. This last result, unveil an important puzzle since Neuroticism is considered an index of sensibility to threats hence, highly Neurotic individuals should be particularly affected in an environment like the Covid-19 period. We further discuss this issue – together with the other main results– in detail in the last section.

The question of whether the lockdown affect the mental health of different individuals in differently way is still very open and compelling. Several studies have been devoted to establishing the effects on different age, gender and ethnicities Banks and Xu (2020); Daly et al. (2020); Davillas and Jones (2020); Etheridge and Spantig (2020); Proto and Quintana-Domeque (2021). However, relatively little attention has been paid to the differential effect of lockdown to different personalities and cognitive skills.

Analyzing the effect of lockdown according to personality and cognitive skills is important at least for two reasons. The first is that it can lead to a more personalized psychological or psychiatric treatment, the second is that understanding how individuals with different personality react to an extreme condition like a lockdown can shed more light on our understanding of personality traits and cognitive skills.

A methodological challenge is represented by unobserved variables that are correlated to both personality and mental health; we cope with this by introducing the family fixed effect that can be introduced thanks to the panel nature of the Understanding Society data. The household fixed effect allows to control for all the time unchanging unobservable at the household levels, like the cultural traits. The study’s results are also robust to the inclusion of controls such as individual education, physical health, income.

## 2 Data

Our main data source is the Covid-19 Survey from the UK Household Longitudinal Study (UKHLS), or Understanding Society. We combine seven waves of the Covid-19 Survey (April, May, June, July, September, November 2020, and January 2021), with Wave 9 main survey (2017-2019), which serves as the baseline for the pre-Covid-19 period (University of Essex, Institute for Social and Economic Research, 2020; University of Essex, Institute for Social and Economic Research, NatCen Social Research, Kantar Public, 2020). This leads to seven panels, each with a during-and pre-Covid-19 period. Each panel is balanced (i.e. contain two observations per respondent) with 6576 data points each.

We apply the longitudinal sampling weights provided in the UKHLS to make inference on the UK population. A key feature of the Covid-19 Survey is that it is longitudinal, enabling individuals to be tracked over the course of the pandemic. In this balanced panel, there are 8772 individuals with data on gender, age and ethnicity (see Table S1 in SI Appendix). We further merged this data with Wave 9 main survey to construct the pre-Covid baseline data, and Wave 3 main survey to include information on personality traits and cognitive skills. At the end of this process we have a total of 5583 individuals and an attrition of about 36% of which about 21% (i.e. determined by the difference between 8772 and 6928) is due to exogenous factors due to the difference in the responders present the different waves, while the other 16% is due to missing data.

While this attrition rate can be considered substantial, it positively compare with previous research using the same data (Daly et al., 2020; Proto and Quintana-Domeque, 2021). This attrition does not significantly bias the panel in terms of gender (*p*-value = 0.35). The final sample is 3.656 years older than the initial one. A main reason is that since Wave 3 main survey, younger individuals have been added to and older individuals have dropped from the Covid-19 Survey. In the final panel the age range is 24–93, while in the initial balanced Covid-19 Study panel, this is 16-96, hence to the extent that we consider this sample as representative of the UK population within the age range of 24–93, the exogenous attrition does nor represent a strong threat to representativity of our sample.

Importantly, there is no indication of attrition based on personality traits, these differences are insignificant between the final sample and initial balanced panel sample and before and after dropping missing values. Furthermore, there is no significant difference in the mental health indicators (GHQ-12) because of the attrition due to the missing data. All that provides support that sample selection bias plays little role on our analysis. All variables included in the regressions and with their statistical descriptions are listed in Table S3 of SI.

### Mental Health

The index of mental health we use is the 12-item General Health Questionnaire, GHQ-12 (Goldberg, 1988). The GHQ-12 is a well-known self-report instrument for evaluating minor psychiatric disorders, which may signal the beginning of serious disorders, where the respondent must report the extent to which 12 symptoms are present in the past few weeks on a Likert scale, we consider the “caseness” formulation ranging from 0 to 12, which represents the number of symptoms felt “more than usual” or “much more than usual” (we present the questionnaire in SI Appendix, Section A.1).

### Big Five Personality Traits

We use the Personality classification based on the big 5 Five-Factor Model, which is the most common classification Goldberg (1993); Digman (1994); Markon et al. (2005). These “Big Five” are: Neuroticism (or Emotional Stability), Extraversion, Conscientiousness, Agreeableness, Openness usually measured through self-report based on the NEO Five-Factor Inventory (see e.g. Costa Jr and McCrae, 2008), with 60 items (12 items per domain). However, scale-development studies have indicated that the Big Five traits can be reliably assessed with a smaller number of items (e.g. Gosling et al., 2003; Benet-Martínez and John, 1998) that can be used in large-scale surveys. The current data are measured with a short 15-item questionnaire (3 per each of the Big-5 trait). A detailed description of the questions are available in SI Appendix, Section A.1. These Data are from Wave 3 of UKHLS main survey datasets (in 2011-13). Borghans et al. (2008) argue that personality traits vary little for individuals aged between 18 and 65. Given that traits and cognitive skills have been measured in 2011–13, we also run a regression by excluding over 60 and under 27s as a robustness check (SI Appendix, Table S5).^2^. In SI Appendix, Table S2, we present the correlation matrix between personality traits, cognitive skills and gender. As it is normally observed, Neuroticism is negatively correlated with all other traits that are otherwise positively correlated withing each other. As it is normally the case, the correlation between Openness and Cognitive skills is positive and rather substantial.

### Control Variables

We use a measure of cognitive skills as a control variable. They have also been measured in Wave 3 main survey of the UKHLS (in 2011–13). We use the 1^*st*^ principal component of all measures provided in the main UKHLS dataset, apart from the self-rated memory (the questions are presented in SI Appendix, Section A.1 (see McFall, 2013, for details).

Moreover, we introduce: job status, household income (in logarithm) missing income (dummy) any long term health condition, month of the interview, age, region, marital status, household size and presence of children. Also these data are from in the UKHLS dataset. The way these variables have been included in the regressions with their statistical descriptions are listed in SI Appendix, Table S3.

## 3 Econometric Models

We have a series of balanced panels with two periods each, so every respondent is recorded twice: once in the pre-Covid-19 wave (i.e. Wave 9 main survey, related to period 2018/19) and once in each of the wave within the Covid-19 period (April, May, June, July, September, November 2020, January 2021). Using this dataset, we estimate the following model for each two-period panel:

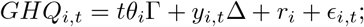

where *i* represents the individual, *t* = 0 indicates the period of Wave 9 main survey, and *t* = 1 denotes each period of the seven waves during the Covid-19 pandemic. *GHQ*_*i,t*_ is the mental health indicator, *θ*_*i*_ is the vector of the time-invariant individual characteristics, including personality traits—our variable of interest, cognitive skills and gender, *y*_*i,t*_ are the time-variant control variables for the each respondent (e.g. income), and *r*_*i*_ are the individual-specific fixed effects. The vector of time invariant characteristics is:

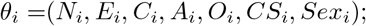

where *N* = Neuroticism, *E* = Extraversion, *C* = Conscientiousness, *A* = Agreeableness, *O* = Openness, *CS* = Cognitive Skills, and *Sex* = Female. The term *tθ*_*i*_Γ represents the interaction of a personality trait and other time-invariant individual characteristics with *t*, which is equal to 1 in Covid-19 period and 0 otherwise. Therefore, some components of vector Γ represent our main the coefficients of interest. *E*_*i,t*_ is an idiosyncratic error assumed, as usual, to be uncorrelated with the regressors. In the regression estimating Equation 3, we cluster the standard errors at the individual levels (i.e. we make the standard assumption, given the above specification of the model with individual fixed effects, that errors are uncorrelated across individuals, but correlated within).

## 4 Result

## 5 Results

The results reported in Table 1 represent a sanity check for our data. We will argue now that the results reported in this table are consistent with the findings in the existing mental health literature. Table 1 presents the correlations between mental health, as measured by GHQ-12 caseness, and some individual characteristics based on the pre-Covid wave of data. We considered 3 different specifications of the model. In the model reported in column 1, we include personality traits as the only regressors. In column 2, we include controls for gender and cognitive skills. In column 3, we further add a number of control variables that will be used for the estimation of our main model that will be presented below.

**Table 1:** Personality and Mental Health in the Pre-Covid-19 Wave

|  | Dep. var. = GHQ12 (0 to 12) |  |  |
| --- | --- | --- | --- |
|  | (1)<br>Wave 9<br>2017–19 | (2)<br>Wave 9<br>2017–19 | (3)<br>Wave 9<br>2017–19 |
| Female |  | 0.312***<br>(0.074) | 0.133*<br>(0.074) |
| Agreeableness | 0.047<br>(0.040) | 0.021<br>(0.041) | 0.025<br>(0.039) |
| Conscientiousness | −0.108***<br>(0.040) | −0.126***<br>(0.040) | −0.073*<br>(0.039) |
| Extraversion | −0.036<br>(0.038) | −0.062<br>(0.038) | −0.066*<br>(0.037) |
| Neuroticism | 0.748***<br>(0.039) | 0.709***<br>(0.041) | 0.610***<br>(0.040) |
| Openness | 0.082**<br>(0.039) | 0.108***<br>(0.040) | 0.083**<br>(0.039) |
| Cognitive ability |  | −0.036<br>(0.036) | −0.002<br>(0.039) |
| *N* | 6,146 | 6,146 | 6,146 |

*Notes* Personality and cognitive skills variables are standardized. In the model estimated in column 3, control for job status, household income, any long term condition, month of the interview, age, region, marital status, household size and presence of children are included in the estimation but omitted. Robust standard errors are in parentheses. \*\*\*, \*\*, and \* denote statistical significance at 0.01, 0.05, and 0.1 levels respectively.

First of all, from columns 2 and 3, we note that female respondents report more symptoms than men. As it is well known, women tend to report higher level of depression and anxiety.^3^ For columns 2 and 3, we do not observe any significant correlation between cognitive skills and mental health: to the best of our knowledge we do not know any systematic link between these two variables. Considering now personality traits, the correlations reported in Table 1 are broadly consistent with the existing literature on mental health and personality. There is strong positive effect of Neuroticism (roughly, an increase in one standard deviation in Neuroticism is associated with 0.7 more symptoms). Past studies also find negative effects of Conscientiousness on mental health deterioration and, to a lower extent, of Extraversion; while Agreeableness is largely uncorrelated with mental health. Table 4 in (Kotov et al., 2010) report these effects on anxiety and depression (see also Klein et al., 2011). Correlations in Table 1 are also consistent with the links observed between personality traits and self-reported subjective wellbeing (or happiness as it is frequently called), with Neuroticism correlating negatively, while Conscientiousness and Extraversion usually correlate positively (Watson and Clark, 1992; Diener and Lucas, 1999). The only exception in the comparison with the existing literature on mental health is the significant correlation between GHQ-12 and Openness that we can observe in Table 1. A possible explanation is that mental health studies as the ones reviewed in Kotov et al. (2010) consider serious disorders, while GHQ-12 is better suited for detecting minor psychiatric disorders (which may signal the beginning of serious disorders). This explanation is supported by the frequently observed negative associations between Openness and subjective wellbeing (Watson and Clark, 1992; Proto and Rustichini, 2015).

It is also important to note that the coefficients reported in Table 1 over Openness, albeit statistically significant, are much smaller than the one we find on Neuroticism.^4^

The top left panel of Fig. 1 presents the evolution of average mental health deterioration, as measured by the increase in GHQ-12 between each wave during Covid-19 and the baseline (2018–19), for all selected respondents from March 2020 to January 2021.

**Figure 1:**
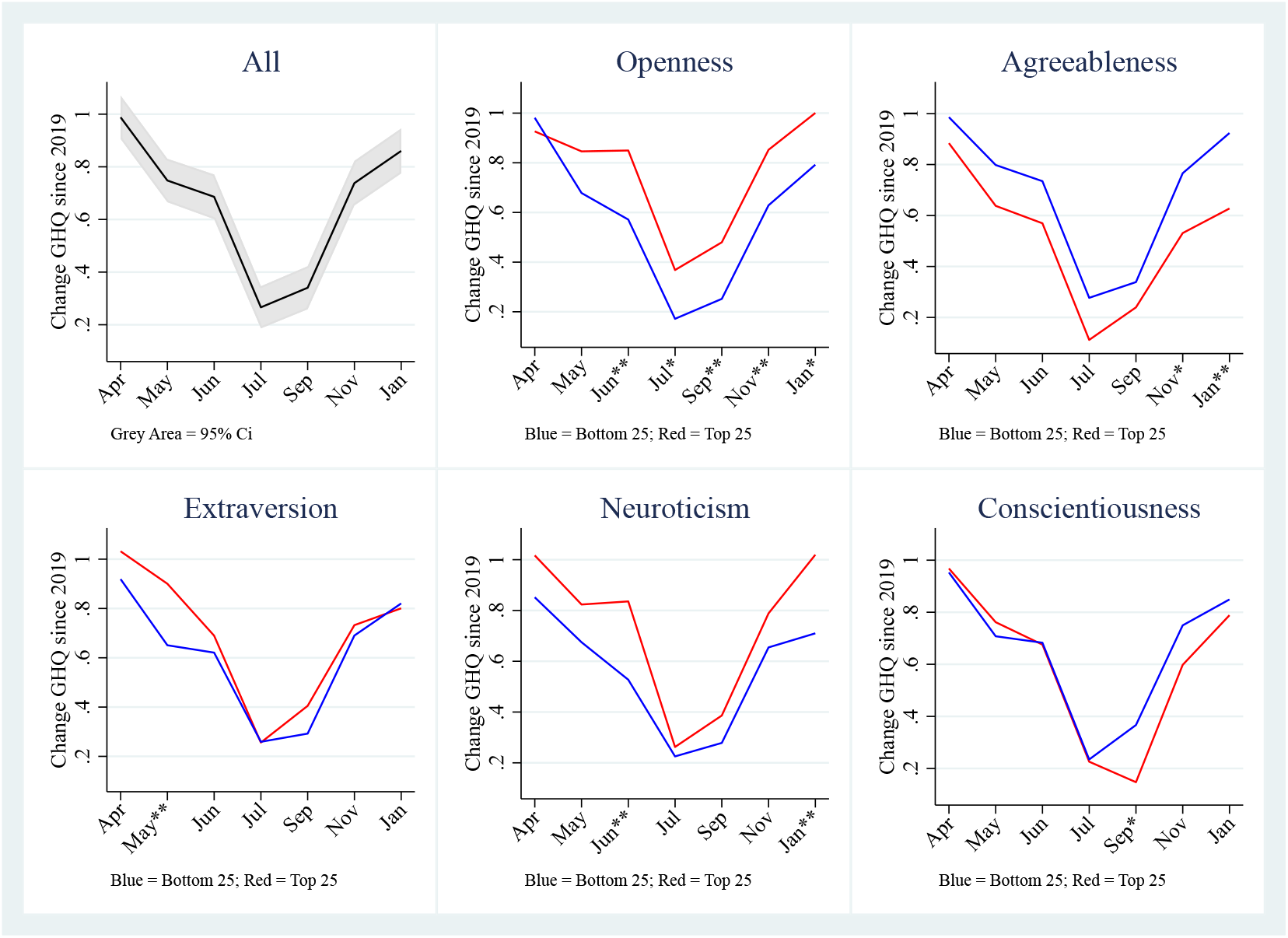
Mental health deterioration in the Covid-19 period, in total and among individuals with different personalty traits. The changes in GHQ-12 represent mental health deterioration between the pre-Covid wave and each wave during the Covid-19 period. The black line in the top left panel represents the overall average, while the other panels report the averages among subjects with the top (red lines) and bottom (blue lines) 25% score in each personality trait. GHQ-12 index is the number of symptoms—up to 12—indicating some form of mental disorders. ** and * next to the months denote statistical significance of the difference between the two lines at 0.05, and 0.1 levels respectively.

We note a timeline of significant Covid-19 restriction policies adopted by the UK government as below. On March 23rd 2020, the Prime Minister announces UK wide lockdown; on May 10th, ‘Stay at home’ becomes ‘stay alert’ and the Prime Minister (PM) sets out lockdown lifting plan; on July 4th, most restrictions are lifted in England. On October 31st, PM announces that England is placed under another national lockdown. On December 2nd, England’s national lockdown comes to an end and is replaced by a strengthened three-tier system. On January 4th 2021, PM announces a third national lockdown for England.

In Figure 1, we observe a V-shaped path of mental health deterioration from April 2020 to January 2021. Figure 1 shows a dramatic rise in GHQ-12 in April of about one unit (i.e. one more symptom per individual) then a decline during late spring and early summer and an increase again in autumn 2020 and January 2021. This path roughly mirrors the evolution of the infections and restrictions. The average mental health deterioration (i.e. average GHQ-12 changes) over the entire period from April 2020 to January 2021 is around 0.66 symptoms (i.e. two out of three respondents experienced one more symptom on average).

The other five panels of Figure 1 present the GHQ-12 evolution for individuals scoring high and low in each personality trait (more precisely, belonging to top and bottom 25% of each personality score). A visual inspections of the five panels reveals clear differences in mental health deterioration for respondents at the top and bottom ends of all five traits. In particular, individuals high in Openness and low in Agreeableness seem to have experienced worse mental health deterioration than their counterparts to the other extremes. Neuroticism seems to affect individuals in the natural direction, i.e. respondents scoring high in Neuroticism experienced worse mental health deterioration than those scoring low. Extraversion seems to have more heavily affected respondents at the beginning of the period, while Conscientiousness in the second half.

The evidence presented in Figure 1 provides a first indication of a differential impact of the Covid-19 period on mental health. There are, however, some potential confounding factors in the relationship between personality and mental health deterioration during the period of analysis. For example, personality can affect the probability of becoming unemployed or lead a lower wage during the Covid-19 period (e.g. Borghans et al., 2008; Proto and Rustichini, 2015) and, in turn, both these factors that can affect the mental health. Therefore, as previously argued in Section 3, we move on to estimate model 3 which controls for such confounding factors, to assess the relationships suggested in Figure 1.

Table 2 presents the estimations of model 3, for each of the seven panels of data consisting of the pre-Covid wave (i.e. Wave 9 main survey) and one of the waves during the Covid-19 period, i.e.: April 2020 (column 1), May (column 2), June (column 3), July (column 4), September (column 5), November(column 6)), and January 2021 (column 7). For ease of visualisation, we present coefficient plots of interaction terms between personality traits and Covid-19 period in SI Appendix, Figure S1.

**Table 2:**
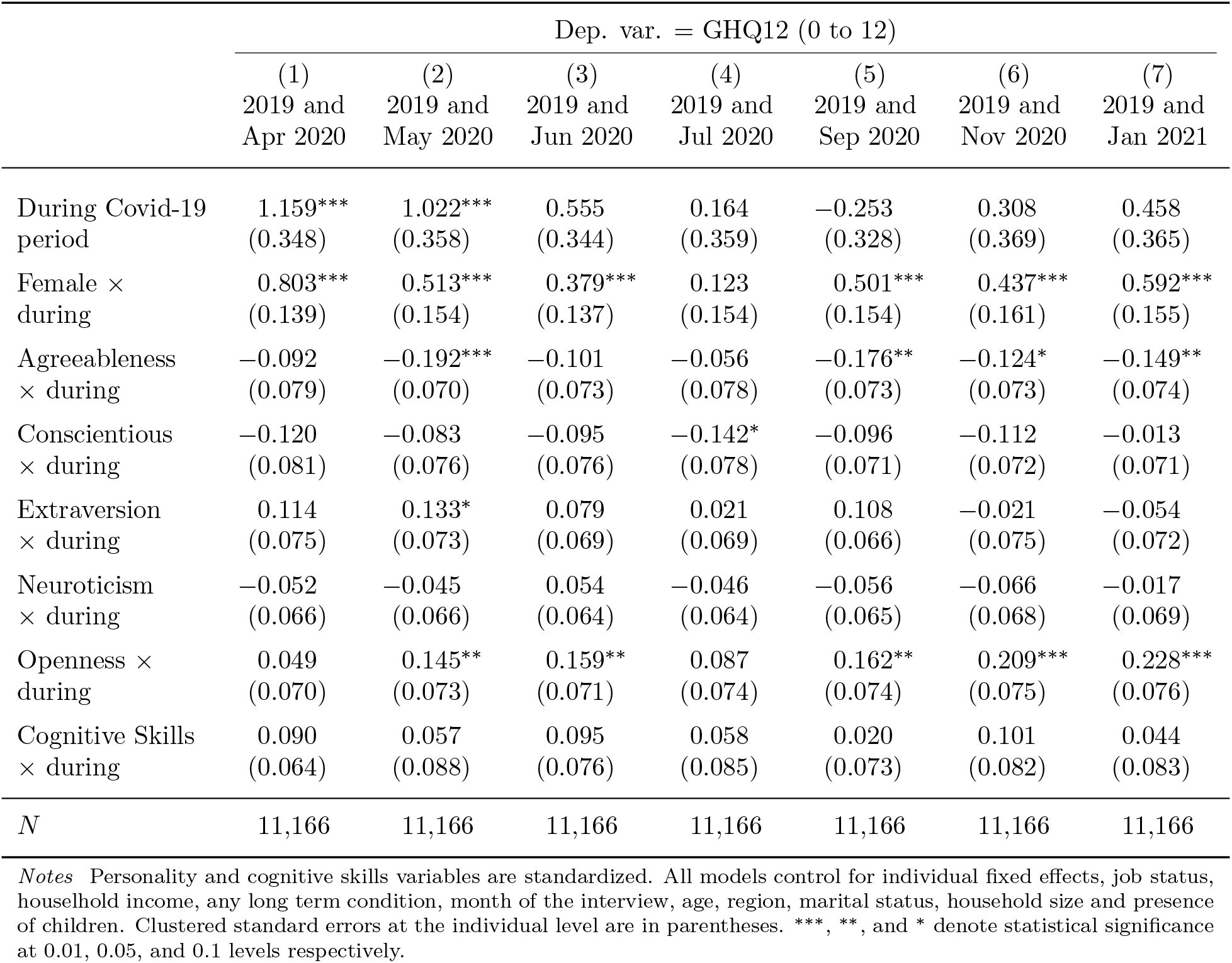
Personality and Mental Health Deterioration During the Covid-19 Period

First of all, we observe in Table 2 that, consistent with existing evidence, female respondents report more symptoms of mental health deterioration than males, during the Covid-19 period (Banks and Xu, 2020; Daly et al., 2020; Davillas and Jones, 2020; Etheridge and Spantig, 2020; Proto and Quintana-Domeque, 2021). Concerning the personality traits, in general we note that some of them significantly predict more mental health deterioration, with a non-negligible magnitude. To have an idea, we notice that a coefficient of about 0.15 implies that one standard deviation in personality increases of 0.15 symptoms on the GHQ-12 measure, i.e. one out of seven respondents reporting one more symptom in the Covid-19 period; and we recall that the average mental health deterioration in the Covid-19 period is about 0.69 more symptoms.

In particular, personalities with low score in Agreeableness and high score in Openness predict more mental health deterioration during the Covid-19 period. The effect of Openness seems to be increasing throughout the period and it is remarkably high in January 2021, where a one standard deviation increase in Openness predicts an increase of 0.23 symptoms on average (see also Figure S1 in SI Appendix). The interaction with Extraversion is weakly significant in the second period, but if we consider the GHQ-12 score (scale 0–36) instead (SI Appendix, Table S6) this becomes strongly significant at 5% level for the second period and marginally significant at 10% for the first and third periods. The interaction with Conscientiousness is weakly significant in the 4^*th*^ wave. Neuroticism is surprisingly insignificant in this specification.^5^

To make sure we are not picking up diverging trends or time effects due to different personality traits, we further run a placebo test, with Wave 9 main survey as the intervention period and Wave 8 as the baseline period (see Appendix SI, Table S7). In this test, we are not able to detect any significant differential effects due to personality traits across these two waves, lending support to the notion that the diverging trends in mental health across different levels of personality traits are specific to the Covid-19 period. We also check for robustness to other psychological factors that might be correlated with personality traits, including optimism, risk attitude, and locus of control (see Appendix SI, Table S8). The results are also qualitatively similar if we omit sampling weights (SI Appendix, Table S9), measure mental health with GHQ12 scale instead of caseness (SI Appendix, Table S10), exclude those over 60s and under 27s (SI Appendix, Table S11), or consider different specifications of model 3 (SI Appendix, Tables S12 and S13).

In Table 3, we report the results of the estimation of model 3 for males and females separately.^6^ Even if some coefficients lose significance in comparison with the estimations presented in Table 2, given the lower power of this test, we note that both Openness and Cognitive Skills (which is insignificant when we consider all together) are particularly strong predictors of mental health deterioration in female respondents.

**Table 3:** Personality and Mental Health Deterioration During the Covid-19 Period for Male and Female

|  | Dep. var. = GHQ12 (0 to 12) |  |  |  |  |  |
| --- | --- | --- | --- | --- | --- | --- |
|  | (1) | (2) | (3) | (4) | (5) | (6) |
|  | 2019 and<br>May 2020<br>Female | 2019 and<br>May 2020<br>Male | 2019 and<br>Jul 2020<br>Female | 2019 and<br>Jul 2020<br>Male | 2019 and<br>Jan 2021<br>Female | 2019 and<br>Jan 2021<br>Male |
| During Covid-19<br>period | 1.656***<br>(0.391) | 0.863<br>(0.530) | -0.121<br>(0.444) | 0.672<br>(0.480) | 1.198**<br>(0.505) | 0.259<br>(0.471) |
| Agreeableness<br>× during | -0.182*<br>(0.103) | -0.199**<br>(0.090) | -0.101<br>(0.126) | -0.028<br>(0.092) | -0.134<br>(0.101) | -0.149<br>(0.107) |
| Conscientious<br>× during | -0.056<br>(0.099) | -0.135<br>(0.116) | -0.152<br>(0.119) | -0.118<br>(0.096) | -0.008<br>(0.096) | -0.051<br>(0.105) |
| Extraversion<br>× during | 0.187*<br>(0.100) | 0.082<br>(0.100) | 0.102<br>(0.099) | -0.056<br>(0.086) | -0.152<br>(0.103) | 0.063<br>(0.097) |
| Neuroticism<br>× during | -0.019<br>(0.089) | -0.080<br>(0.101) | -0.030<br>(0.087) | -0.040<br>(0.093) | 0.013<br>(0.093) | -0.022<br>(0.102) |
| Openness ×<br>during | 0.132<br>(0.094) | 0.158<br>(0.115) | 0.050<br>(0.096) | 0.129<br>(0.111) | 0.284***<br>(0.091) | 0.162<br>(0.126) |
| Cognitive Skills<br>× during | 0.198**<br>(0.084) | -0.130<br>(0.158) | 0.233***<br>(0.084) | -0.171<br>(0.152) | 0.136<br>(0.085) | -0.074<br>(0.135) |
| *N* | 8,806 | 7,943 | 8,806 | 7,943 | 8,806 | 7,943 |

*Notes* Personality and cognitive skills variables are standardized. All models control for individual fixed effects, job status, household income, any long term condition, month of the interview, age, region, marital status, household size and presence of children. Clustered standard errors at the individual level are in parentheses. \*\*\*, \*\*, and \* denote statistical significance at 0.01, 0.05, and 0.1 levels respectively.

To summarize our empirical findings, we can say that *During the Covid-19 period Agreeableness is a negative predictor of Mental Health deterioration; while Openness and, to a lower extent, Extraversion are positive predictors; Neuroticism is surprisingly insignificant in all specifications of the model. In female respondents, Cognitive Skills and Openness are particularly strong predictors of mental health deterioration*

## 6 Discussion

Our results show that Openness is a strong predictor of mental health deterioration during the pandemic period. Openness is the trait that reflects preferences for exploration and new experiences (DeYoung et al., 2005; DeYoung and Gray, 2009), in fact this trait is often called “Openness to Experience”. The pandemic period is characterized by several constraints that limit the capacity of making new experience or seeking new sensations, and the fact that Openness is positively associated with mental health deterioration reflect this view. Furthermore, Openness is among the Big Five trait the one that is more consistently positively associated with intelligence (as we can observe in SI Appendix, Table S2 for our data as well), in fact Openness is sometime referred as “Intellect”. Cognitive skills like fluid intelligence and working memory seem to be related primarily to the aspect of Openness/Intellect that can be described as Intellect, which can be separated by the artistic and contemplative traits that characterize the Openness aspect (DeYoung et al., 2005, 2007). In our main analysis we introduce cognitive skills as a regressor together with Openness, hence we can separately analyze the two aspects of Openness and Intellect. Interestingly Openness and Cognitive skills are particularly strong negative predictors of mental health for women, while there is no significant effect of cognitive skills for men.

Agreeableness reflects a tendency toward the maintenance of social stability, for this reason an individual with a more Agreeable personality can cope better in the constrained environment following the lockdown (DeYoung and Gray, 2009). However, at the same time, individuals scoring high in Agreeableness should have a general altruistic tendency, and tend to be interested in and considerate of others’ needs and feelings. In the pandemic, the knowledge that other people, either within the family or outside are suffering for various reasons, can negatively affect individuals with a more agreeable personality. Our evidence suggest that the first effect is stronger than the second.

Extraversion is, generally speaking, a trait related to sensitivity to social rewards (e.g. Depue and Morrone-Strupinsky, 2005). Therefore, in an environment where social contacts are restricted, it is natural to expect that extrovert individuals are particularly negatively affected. The fact that this seems to be true only in the first part of Covid-19 period might be due to the fact that Extravert responders managed to adapt to this situation, perhaps by using the social media platforms.

Neuroticism is linked to higher sensitivity to negative emotions like anger, hostility or depression. For this reason Neuroticism is associate with sensibility to negative outcomes and threats (DeYoung and Gray, 2009) that should be pervasive during the current pandemic. Surprisingly, on our data we find only a weak evidence of this. A possible answer is that, given that as we can observe from Table 1. Neuroticism is a strong negative predictor of mental health deterioration in general, and individuals with highly neurotic personality have normally experienced several negative shocks in the course of their lives, hence there might be a sort of habituation effect playing. Another possibility is that each individual does not normally experience too many symptoms of mental health deterioration, hence responders with an highly Neurotic personality cannot experience more symptoms than what they experienced before the pandemic period.

Conscientiousness reflects a tendency to maintain motivational stability. For this reason, a conscientious individual can overcome better the practical constraints and manage better the negative feelings due to the pandemics. On the other hand, conscientious individuals have preferences to make long-term ambitious plans, something impossible to achieve in an highly uncertain environment, hence there is no reason to expect a positive or negative effect.

## Data Availability

The University of Essex Ethics Committee has approved all data collection on Understanding Society main study and innovation panel waves, including asking consent for all data linkages except to health records. Requesting consent for health record linkage was approved at Wave 1 by the National Research Ethics Service (NRES) Oxfordshire REC A (08/H0604/124), at BHPS Wave 18 by the NRES Royal Free Hospital & Medical School (08/H0720/60) and at Wave 4 by NRES Southampton REC A (11/SC/0274)

## Acknowledgements

Understanding Society is an initiative funded by the Economic and Social Research Council and various Government Departments, with scientific leadership by the Institute for Social and Economic Research, University of Essex, and survey delivery by NatCen Social Research and Kantar Public. The research data are distributed by the UK Data Service. The authors thank several colleagues for discussions on this and related research, in particular Dimitris Christellis, Dietmar Fehr, Vittal Katikireddi, Rajesh Ramachandran, Andis Sofianos and seminar participants at the University of Heidelberg. Responsibility for errors and confusion is ours.

### A SI Appendix

#### A.1 Definition of Key Variables

This section provides details on how these key variables are defined: GHQ-12, Big Five personality, and cognitive skills.

##### GHQ-12 Questionnaire

The GHQ module contains the following questions for the GHQ-12 Questionnaire:

1. scghqa [GHQ: concentration] The next questions are about how you have been feeling over the last few weeks. Have you recently been able to concentrate on whatever you’re doing? 1. Better than usual 2. Same as usual 3. Less than usual 4. Much less than usual
2. scghqb [GHQ: loss of sleep] Have you recently lost much sleep over worry? 1. Not at all 2. No more than usual 3. Rather more than usual 4. Much more than usual
3. scghqc [GHQ: playing a useful role] Have you recently felt that you were playing a useful part in things? 1. More so than usual 2. Same as usual 3. Less so than usual 4. Much less than usual
4. scghqd [GHQ: capable of making decisions] Have you recently felt capable of making decisions about things? 1. More so than usual 2. Same as usual 3. Less so than usual 4. Much less capable
5. scghqe [GHQ: constantly under strain] Have you recently felt constantly under strain? 1. Not at all 2. No more than usual 3. Rather more than usual 4. Much more than usual
6. scghqf [GHQ: problem overcoming difficulties] Have you recently felt you couldn’t overcome your difficulties? 1. Not at all 2. No more than usual 3. Rather more than usual 4. Much more than usual
7. scghqg [GHQ: enjoy day-to-day activities] Have you recently been able to enjoy your normal day-to-day activities? 1. More so than usual 2. Same as usual 3. Less so than usual 4. Much less than usual
8. scghqh [GHQ: ability to face problems] Have you recently been able to face up to problems? 1. More so than usual 2. Same as usual 3. Less able than usual 4. Much less able
9. scghqi [GHQ: unhappy or depressed] Have you recently been feeling unhappy or depressed? 1. Not at all 2. No more than usual 3. Rather more than usual 4. Much more than usual
10. scghqj [GHQ: losing confidence] Have you recently been losing confidence in yourself? 1. Not at all 2. No more than usual 3. Rather more than usual 4. Much more than usual
11. scghqk [GHQ: believe worthless] Have you recently been thinking of yourself as a worthless person? 1. Not at all 2. No more than usual 3. Rather more than usual 4. Much more than usual
12. scghql [GHQ: general happiness] Have you recently been feeling reasonably happy, all things considered? 1. More so than usual 2. About the same as usual 3. Less so than usual 4. Much less than usual The GHQ-12 range goes from 0-36. This range is obtained by subtracting 1 to the values given in each question. Thus, the values in each question are re-coded from 1-4 to 0-3.

##### The “Big Five” in the Understanding Society Survey

The big five personality traits are Agreeableness (A), Conscientiousness (C), Extraversion (E), Neuroticism (N), Openness (O). They are assessed with the following questions:

I see myself as someone who:

1. (A) Is sometimes rude to others (*reverse-scored*).
2. (C) Does a thorough job.
3. (E) Is talkative.
4. (N) Worries a lot.
5. (O) Is original, comes up with new ideas.
6. (A) Has a forgiving nature.
7. (C) Tends to be lazy (*reverse-scored*).
8. (E) Is outgoing, sociable.
9. (N) Gets nervous easily.
10. (O) Values artistic, aesthetic experiences.
11. (A) Is considerate and kind to almost everyone.
12. (C) Does things efficiently.
13. (E) Is reserved (*reverse-scored*).
14. (N) Is relaxed, handles stress well (*reverse-scored*).
15. (O) Has an active imagination.

##### The Cognitive Skills in the Understanding Society Survey

###### Episodic Memory or Memory

“The computer will now read a set of 10 words. I would like you to remember as many as you can. We have purposely made the list long so it will be difficult for anyone to remember all the words. Most people remember just a few. Please listen carefully to the set of words as they cannot be repeated. When it has finished, I will ask you to recall aloud as many of the words as you can, in any order. Is this clear? Now please tell me the words you can remember.” Respondents give the words in any order. The interviewer codes each correct response… For the delayed word recall test, after the Number Series test, respondents were again asked to remember the words from the list. The interviewer codes each correct response.

###### Working Memory or Serial 7 Subtraction

“Now let’s try some subtraction of numbers. One hundred minus 7 equals what?’ [Interviewer records the number.] ‘And take 7 away from that?’ [records number] ‘And take 7 away from that.’ The respondent gives numeric answers for successive trials, five in all.”

###### Verbal Fluency

“Now, I would like you to name as many animals as you can. You have one-minute, so name them as quickly as possible. We will begin when you say the first animal. If you are unsure of anything please ask me now as I am unable to answer questions once the minute starts”

**Table S.1:**
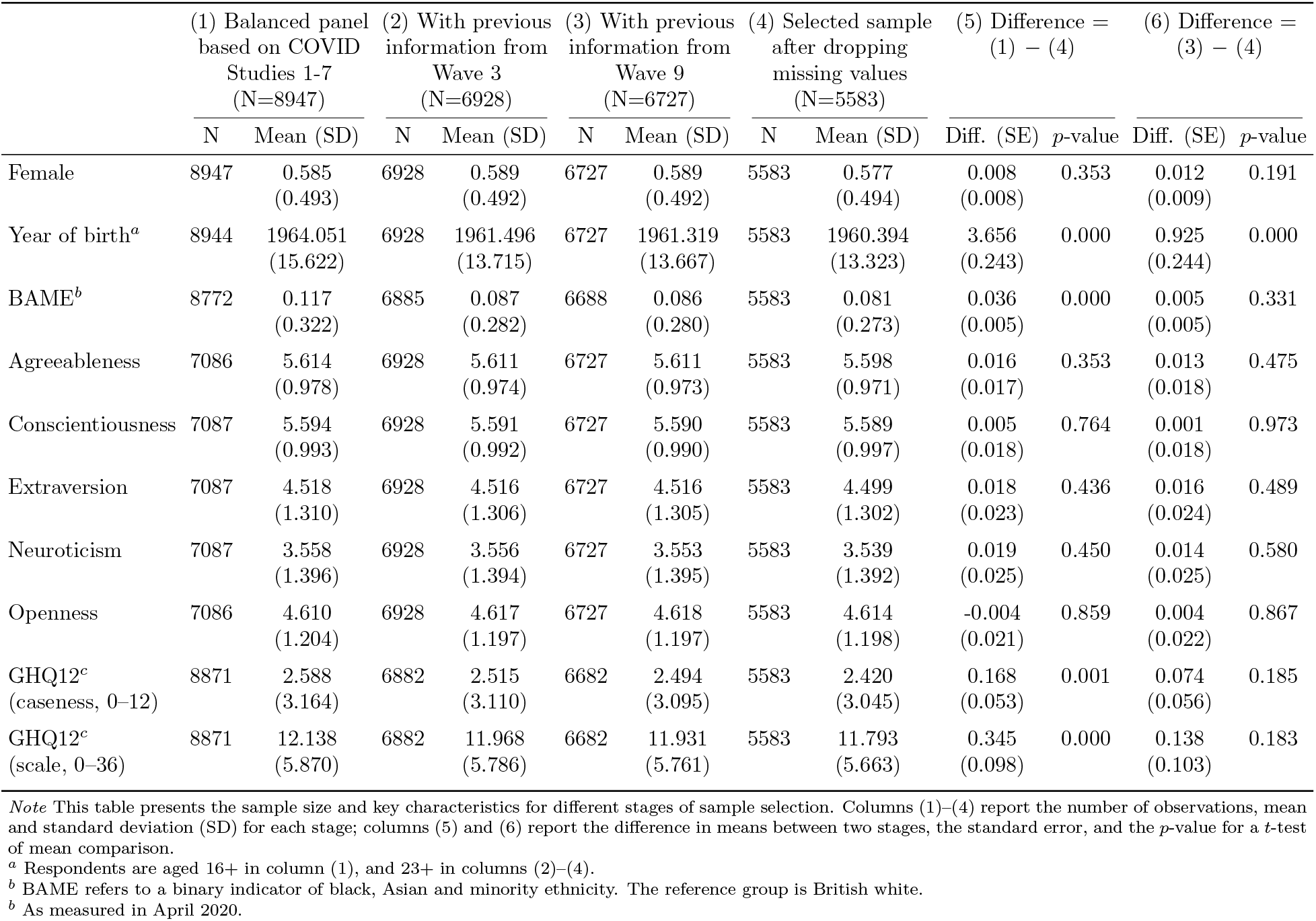
Sample size and key characteristics at different stages of sample selection

###### Problem Solving or Numerical Ability

“Next I would like to ask you some questions to understand how people use numbers in everyday life. If CATI, the interviewer added, You might want to have a pencil and paper handy to help you answer the following items The measure of numeric ability asks respondents up to five questions that are graded in complexity (Table 2 at page 14 of McFall, 2013, displays the questions and how they are administered)”

###### Fluid Reasoning or Number Series

Individuals are randomly assigned to Set 1 or Set 2 (of items) within households. For this test, respondents use a pencil and paper to write down the number sequences as read by the interviewer. The number series consists of several numbers with a blank number in the series. The respondent will be asked which number goes in the blank (see page 11 of McFall, 2013, for more details)

#### A.2 Additional Results

This section provides additional tables that are not included in the main text.

**Table S.2:**
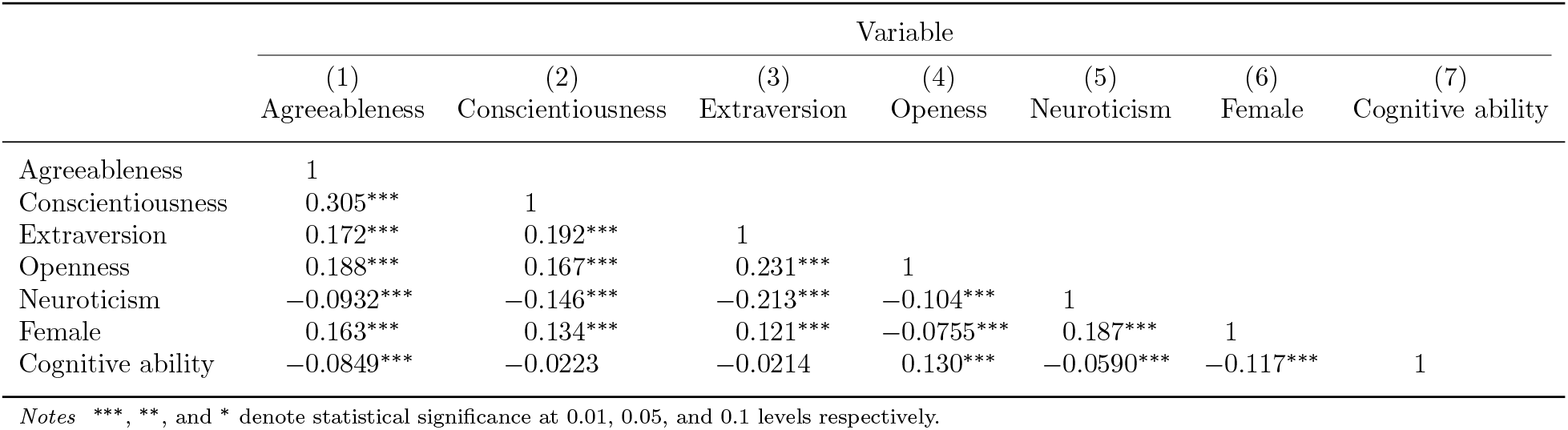
Correlation matrix

**Table S.3:**
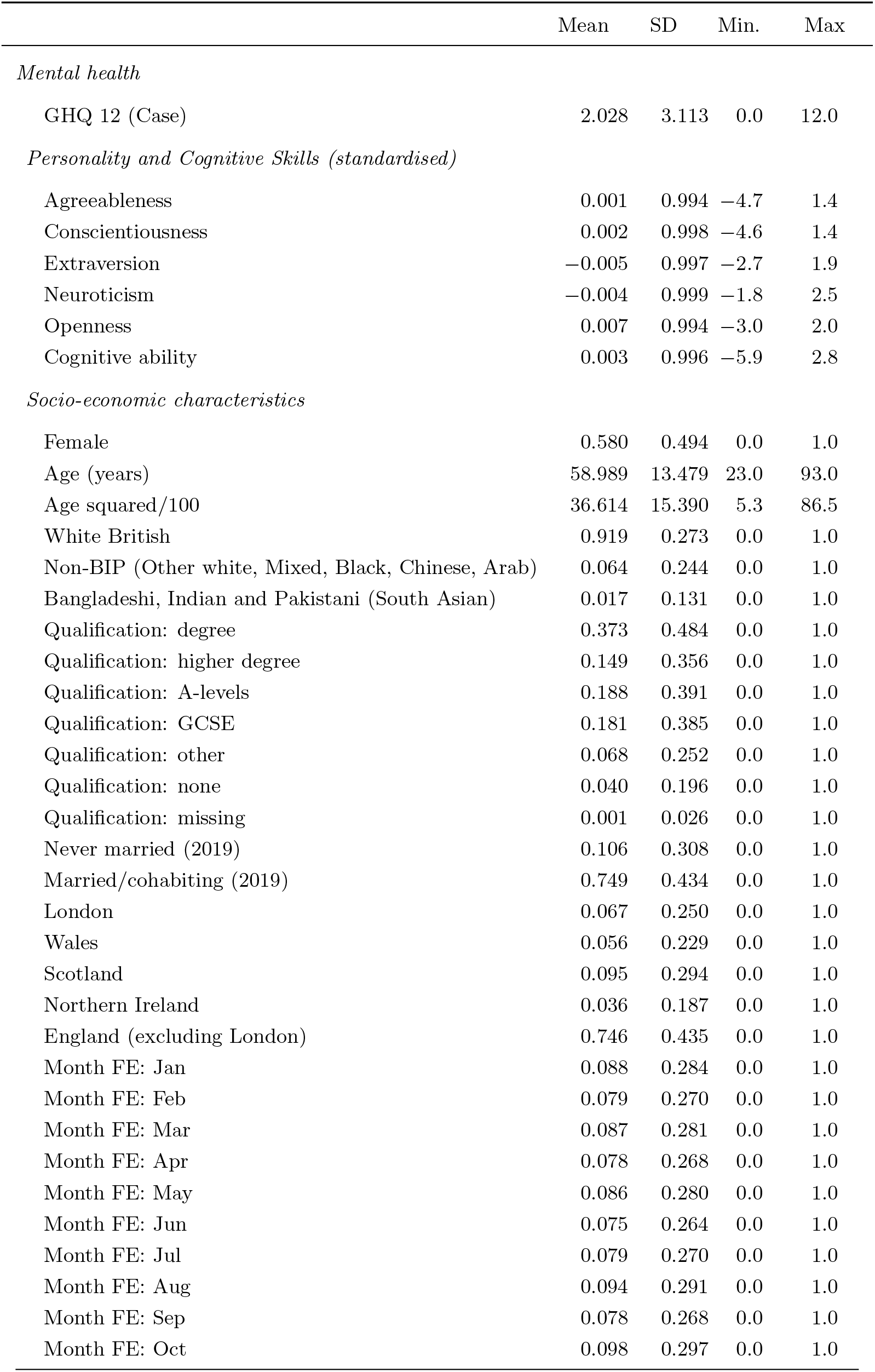
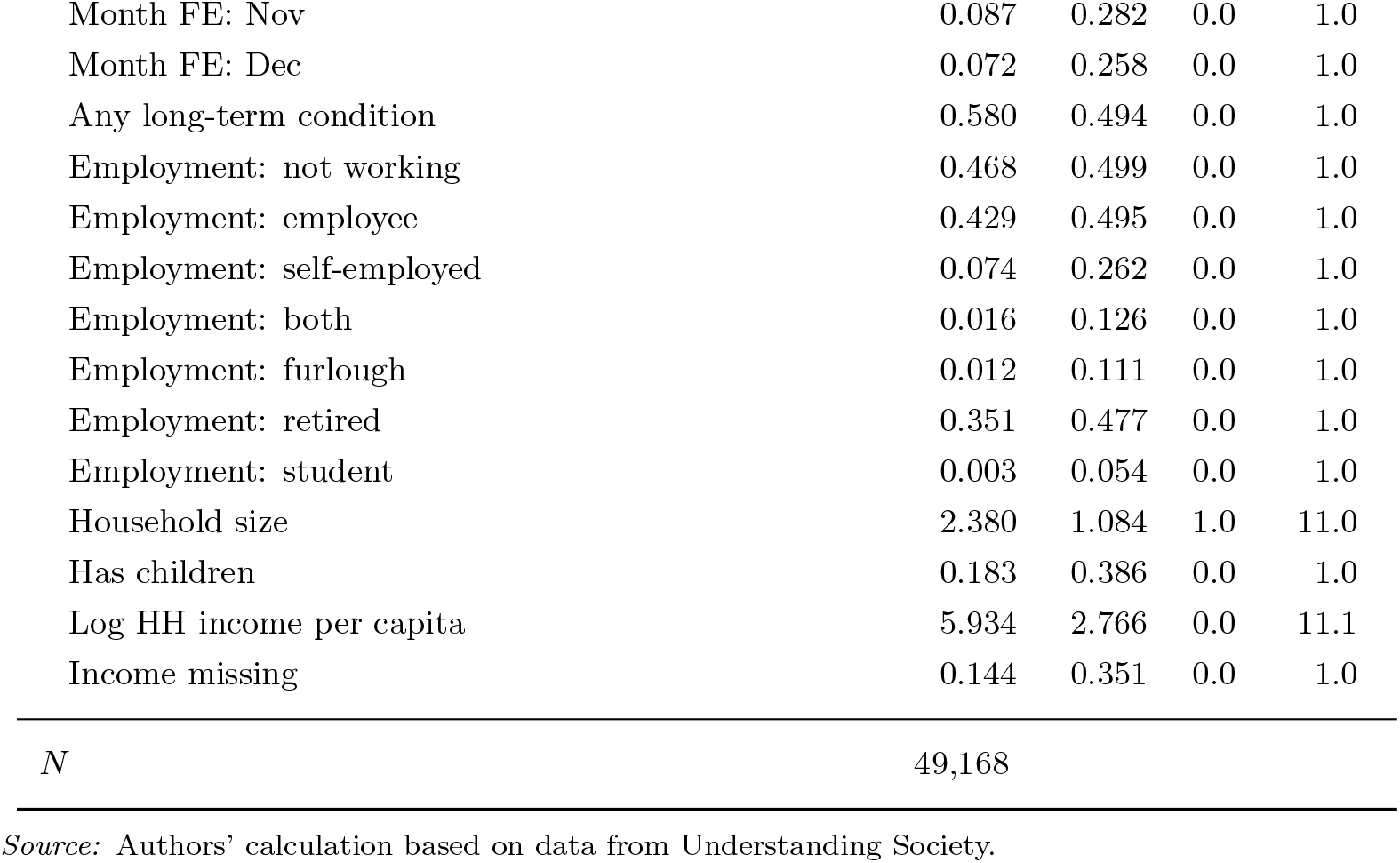
Summary statistics

|  | Mean | SD | Min. | Max |
| --- | --- | --- | --- | --- |
| *Mental health* |  |  |  |  |
| GHQ 12 (Case) | 2.028 | 3.113 | 0.0 | 12.0 |
| *Personality and Cognitive Skills (standardised)* |  |  |  |  |
| Agreeableness | 0.001 | 0.994 | -4.7 | 1.4 |
| Conscientiousness | 0.002 | 0.998 | -4.6 | 1.4 |
| Extraversion | -0.005 | 0.997 | -2.7 | 1.9 |
| Neuroticism | -0.004 | 0.999 | -1.8 | 2.5 |
| Openness | 0.007 | 0.994 | -3.0 | 2.0 |
| Cognitive ability | 0.003 | 0.996 | -5.9 | 2.8 |
| *Socio-economic characteristics* |  |  |  |  |
| Female | 0.580 | 0.494 | 0.0 | 1.0 |
| Age (years) | 58.989 | 13.479 | 23.0 | 93.0 |
| Age squared/100 | 36.614 | 15.390 | 5.3 | 86.5 |
| White British | 0.919 | 0.273 | 0.0 | 1.0 |
| Non-BIP (Other white, Mixed, Black, Chinese, Arab) | 0.064 | 0.244 | 0.0 | 1.0 |
| Bangladeshi, Indian and Pakistani (South Asian) | 0.017 | 0.131 | 0.0 | 1.0 |
| Qualification: degree | 0.373 | 0.484 | 0.0 | 1.0 |
| Qualification: higher degree | 0.149 | 0.356 | 0.0 | 1.0 |
| Qualification: A-levels | 0.188 | 0.391 | 0.0 | 1.0 |
| Qualification: GCSE | 0.181 | 0.385 | 0.0 | 1.0 |
| Qualification: other | 0.068 | 0.252 | 0.0 | 1.0 |
| Qualification: none | 0.040 | 0.196 | 0.0 | 1.0 |
| Qualification: missing | 0.001 | 0.026 | 0.0 | 1.0 |
| Never married (2019) | 0.106 | 0.308 | 0.0 | 1.0 |
| Married/cohabiting (2019) | 0.749 | 0.434 | 0.0 | 1.0 |
| London | 0.067 | 0.250 | 0.0 | 1.0 |
| Wales | 0.056 | 0.229 | 0.0 | 1.0 |
| Scotland | 0.095 | 0.294 | 0.0 | 1.0 |
| Northern Ireland | 0.036 | 0.187 | 0.0 | 1.0 |
| England (excluding London) | 0.746 | 0.435 | 0.0 | 1.0 |
| Month FE: Jan | 0.088 | 0.284 | 0.0 | 1.0 |
| Month FE: Feb | 0.079 | 0.270 | 0.0 | 1.0 |
| Month FE: Mar | 0.087 | 0.281 | 0.0 | 1.0 |
| Month FE: Apr | 0.078 | 0.268 | 0.0 | 1.0 |
| Month FE: May | 0.086 | 0.280 | 0.0 | 1.0 |
| Month FE: Jun | 0.075 | 0.264 | 0.0 | 1.0 |
| Month FE: Jul | 0.079 | 0.270 | 0.0 | 1.0 |
| Month FE: Aug | 0.094 | 0.291 | 0.0 | 1.0 |
| Month FE: Sep | 0.078 | 0.268 | 0.0 | 1.0 |
| Month FE: Oct | 0.098 | 0.297 | 0.0 | 1.0 |
| Month FE: Nov | 0.087 | 0.282 | 0.0 | 1.0 |
| Month FE: Dec | 0.072 | 0.258 | 0.0 | 1.0 |
| Any long-term condition | 0.580 | 0.494 | 0.0 | 1.0 |
| Employment: not working | 0.468 | 0.499 | 0.0 | 1.0 |
| Employment: employee | 0.429 | 0.495 | 0.0 | 1.0 |
| Employment: self-employed | 0.074 | 0.262 | 0.0 | 1.0 |
| Employment: both | 0.016 | 0.126 | 0.0 | 1.0 |
| Employment: furlough | 0.012 | 0.111 | 0.0 | 1.0 |
| Employment: retired | 0.351 | 0.477 | 0.0 | 1.0 |
| Employment: student | 0.003 | 0.054 | 0.0 | 1.0 |
| Household size | 2.380 | 1.084 | 1.0 | 11.0 |
| Has children | 0.183 | 0.386 | 0.0 | 1.0 |
| Log HH income per capita | 5.934 | 2.766 | 0.0 | 11.1 |
| Income missing | 0.144 | 0.351 | 0.0 | 1.0 |
| *N* | 49,168 |  |  |  |

*Source:* Authors' calculation based on data from Understanding Society.

**Table S.4:** Personality and mental health in each wave

|  | Dep. var. = GHQ12 (from 0 to 12) |  |  |  |  |  |  |  |
| --- | --- | --- | --- | --- | --- | --- | --- | --- |
|  | (1)<br>2017-19 | (2)<br>Apr 2020 | (3)<br>May 2020 | (4)<br>Jun 2020 | (5)<br>Jul 2020 | (6)<br>Sep 2020 | (7)<br>Nov 2020 | (8)<br>Jan 2021 |
| Female | 0.133*<br>(0.074) | 0.863***<br>(0.079) | 0.691***<br>(0.080) | 0.548***<br>(0.085) | 0.392***<br>(0.079) | 0.562***<br>(0.080) | 0.643***<br>(0.087) | 0.691***<br>(0.086) |
| Agreeableness | 0.025<br>(0.039) | 0.004<br>(0.042) | -0.043<br>(0.043) | -0.040<br>(0.045) | 0.024<br>(0.042) | 0.034<br>(0.043) | -0.029<br>(0.045) | -0.066<br>(0.045) |
| Conscientiousness | -0.073*<br>(0.039) | -0.071*<br>(0.041) | -0.095**<br>(0.042) | -0.120***<br>(0.044) | -0.108***<br>(0.041) | -0.169***<br>(0.042) | -0.147***<br>(0.044) | -0.107**<br>(0.045) |
| Extraversion | -0.066*<br>(0.037) | -0.024<br>(0.041) | 0.012<br>(0.041) | -0.027<br>(0.042) | -0.072*<br>(0.040) | -0.032<br>(0.041) | -0.050<br>(0.044) | -0.072*<br>(0.043) |
| Neuroticism | 0.610***<br>(0.040) | 0.632***<br>(0.042) | 0.642***<br>(0.043) | 0.703***<br>(0.045) | 0.600***<br>(0.042) | 0.621***<br>(0.042) | 0.637***<br>(0.045) | 0.689***<br>(0.046) |
| Openness | 0.083**<br>(0.039) | 0.108***<br>(0.042) | 0.165***<br>(0.042) | 0.227***<br>(0.045) | 0.170***<br>(0.042) | 0.193***<br>(0.042) | 0.201***<br>(0.044) | 0.219***<br>(0.045) |
| Cognitive ability | -0.002<br>(0.039) | 0.008<br>(0.042) | 0.032<br>(0.042) | 0.024<br>(0.045) | 0.031<br>(0.041) | 0.015<br>(0.043) | 0.023<br>(0.045) | -0.005<br>(0.046) |
| *N* | 6,146 | 6,146 | 6,146 | 6,146 | 6,146 | 6,146 | 6,146 | 6,146 |

*Notes* Personality and cognitive skills variables are standardized. All models control for job status, household income, any long term condition, month of the interview, age, region, marital status, household size and presence of children. Robust standard errors are in parentheses. \*\*\*, \*\*, and \* denote statistical significance at 0.01, 0.05, and 0.1 levels respectively.

**Figure S.1:**
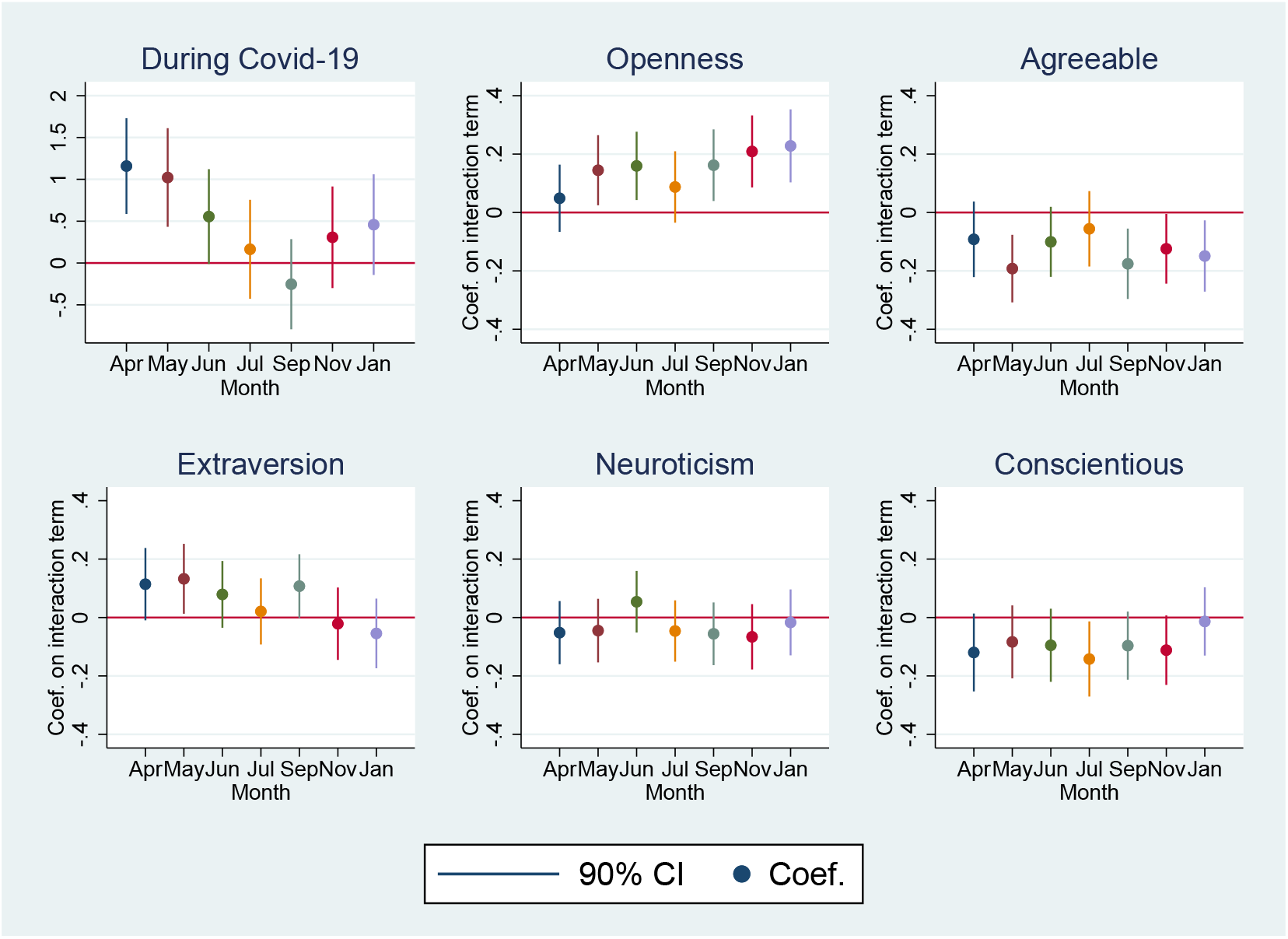
Coefficient plots of the differential effects of Covid-19 period by personality traits

**Table S.5:** Personality and mental health in each wave, excluding over 60s and under 27s

|  | Dep. var. = GHQ12 (from 0 to 12) |  |  |  |  |  |  |  |
| --- | --- | --- | --- | --- | --- | --- | --- | --- |
|  | (1)<br>2018/19 | (2)<br>Apr 2020 | (3)<br>May 2020 | (4)<br>Jun 2020 | (5)<br>Jul 2020 | (6)<br>Sep 2020 | (7)<br>Nov 2020 |  |
| Female | 0.132<br>(0.118) | 1.044***<br>(0.122) | 0.768***<br>(0.127) | 0.656***<br>(0.136) | 0.334**<br>(0.130) | 0.642***<br>(0.129) | 0.792***<br>(0.137) | 0.743***<br>(0.136) |
| Agreeableness | 0.013<br>(0.059) | -0.015<br>(0.062) | -0.082<br>(0.064) | -0.075<br>(0.067) | 0.029<br>(0.063) | 0.052<br>(0.064) | -0.077<br>(0.067) | -0.121*<br>(0.066) |
| Conscientiousness | -0.136**<br>(0.063) | -0.068<br>(0.063) | -0.156**<br>(0.068) | -0.181**<br>(0.071) | -0.119*<br>(0.068) | -0.256***<br>(0.068) | -0.231***<br>(0.069) | -0.132*<br>(0.071) |
| Extraversion | -0.102*<br>(0.060) | 0.010<br>(0.062) | 0.059<br>(0.067) | -0.037<br>(0.067) | -0.058<br>(0.065) | -0.032<br>(0.066) | -0.087<br>(0.071) | -0.075<br>(0.069) |
| Neuroticism | 0.677***<br>(0.061) | 0.617***<br>(0.063) | 0.657***<br>(0.067) | 0.715***<br>(0.068) | 0.649***<br>(0.067) | 0.687***<br>(0.066) | 0.665***<br>(0.070) | 0.708***<br>(0.070) |
| Openness | 0.106*<br>(0.063) | 0.181***<br>(0.062) | 0.267***<br>(0.067) | 0.380***<br>(0.070) | 0.240***<br>(0.066) | 0.305***<br>(0.067) | 0.331***<br>(0.067) | 0.290***<br>(0.070) |
| Cognitive ability | 0.049<br>(0.060) | 0.073<br>(0.064) | 0.041<br>(0.066) | 0.054<br>(0.069) | 0.064<br>(0.065) | 0.008<br>(0.066) | 0.076<br>(0.069) | 0.038<br>(0.070) |
| *N* | 3,185 | 3,040 | 3,040 | 3,040 | 3,040 | 3,040 | 3,040 | 3,040 |

*Notes* Personality and cognitive skills variables are standardized. All models control for job status, household income, any long term condition, month of the interview, age, region, marital status, household size and presence of children. Robust standard errors are in parentheses. \*\*\*, \*\*, and \* denote statistical significance at 0.01, 0.05, and 0.1 levels respectively.

**Table S.6:** Personality and mental health in each wave

|  | Dep. var. = GHQ36 (from 0 to 36) |  |  |  |  |  |  |  |
| --- | --- | --- | --- | --- | --- | --- | --- | --- |
|  | (1)<br>2018/19 | (2)<br>Apr 2020 | (3)<br>May 2020 | (4)<br>Jun 2020 | (5)<br>Jul 2020 | (6)<br>Sep 2020 | (7)<br>Nov 2020 | (8)<br>Jan 2021 |
| Female | 0.303**<br>(0.129) | 1.485***<br>(0.145) | 1.090***<br>(0.143) | 0.900***<br>(0.148) | 0.802***<br>(0.135) | 0.935***<br>(0.137) | 1.071***<br>(0.145) | 1.169***<br>(0.146) |
| Agreeableness | 0.029<br>(0.071) | -0.006<br>(0.079) | -0.037<br>(0.078) | -0.069<br>(0.081) | -0.003<br>(0.072) | 0.050<br>(0.074) | -0.015<br>(0.075) | -0.107<br>(0.079) |
| Conscientiousness | -0.209***<br>(0.067) | -0.217***<br>(0.075) | -0.276***<br>(0.074) | -0.268***<br>(0.077) | -0.250***<br>(0.070) | -0.357***<br>(0.071) | -0.320***<br>(0.074) | -0.266***<br>(0.076) |
| Extraversion | -0.161**<br>(0.064) | -0.114<br>(0.076) | -0.009<br>(0.075) | -0.099<br>(0.074) | -0.174**<br>(0.070) | -0.147**<br>(0.069) | -0.159**<br>(0.074) | -0.173**<br>(0.074) |
| Neuroticism | 1.446***<br>(0.067) | 1.467***<br>(0.077) | 1.483***<br>(0.077) | 1.583***<br>(0.078) | 1.421***<br>(0.072) | 1.431***<br>(0.072) | 1.516***<br>(0.076) | 1.537***<br>(0.078) |
| Openness | 0.044<br>(0.069) | 0.100<br>(0.078) | 0.181**<br>(0.076) | 0.282***<br>(0.078) | 0.195***<br>(0.072) | 0.263***<br>(0.073) | 0.264***<br>(0.077) | 0.282***<br>(0.078) |
| Cognitive ability | -0.026<br>(0.069) | -0.104<br>(0.078) | -0.032<br>(0.076) | -0.044<br>(0.080) | 0.061<br>(0.072) | -0.023<br>(0.073) | -0.017<br>(0.077) | -0.088<br>(0.079) |
| *N* | 6,146 | 6,146 | 6,146 | 6,146 | 6,146 | 6,146 | 6,146 | 6,146 |

*Notes* Personality and cognitive skills variables are standardized. All models control for job status, household income, any long term condition, month of the interview, age, region, marital status, household size and presence of children. Robust standard errors are in parentheses. \*\*\*, \*\*, and \* denote statistical significance at 0.01, 0.05, and 0.1 levels respectively.

**Table S.7:** Placebo test using Wave 8 Main Survey as baseline and Wave 9 Main Survey as intervention year

| Dep. var. = GHQ12 (0 to 12),<br>baseline = Wave 8 main survey,<br>intervention = Wave 9 main survey |  |
| --- | --- |
| Wave 9 | 0.093<br>(0.144) |
| Female ×<br>during Wave 9 | −0.091<br>(0.132) |
| Agreeableness<br>× during | −0.053<br>(0.064) |
| Conscientious<br>× during | 0.057<br>(0.067) |
| Extraversion<br>× during | 0.035<br>(0.062) |
| Neuroticism<br>× during | 0.021<br>(0.066) |
| Openness ×<br>during | 0.003<br>(0.060) |
| Cognitive Skills<br>× during | 0.009<br>(0.061) |
| *N* | 11,301 |

*Notes* Personality and cognitive skills variables are standardized. All models control for individual fixed effects, job status, household income, any long term condition, month of the interview, age, region, marital status, household size and presence of children. Clustered standard errors at the individual level are in parentheses. \*\*\*, \*\*, and \* denote statistical significance at 0.01, 0.05, and 0.1 levels respectively.

**Table S.8:** Robustness check on other psychological factors

|  | Dep. var. = GHQ12 (0 to 12) |  |  |  |  |  |  |
| --- | --- | --- | --- | --- | --- | --- | --- |
|  | (1) | (2) | (3) | (4) | (5) | (6) | (7) |
|  | 2019 and<br>Apr 2020 | 2019 and<br>May 2020 | 2019 and<br>Jun 2020 | 2019 and<br>Jul 2020 | 2019 and<br>Sep 2020 | 2019 and<br>Nov 2020 | 2019 and<br>Jan 2021 |
| During Covid-19<br>period | 0.536<br>(0.625) | 1.375*<br>(0.706) | 0.818<br>(0.594) | 0.693<br>(0.723) | 0.166<br>(0.720) | 0.793<br>(0.760) | 1.380*<br>(0.750) |
| Female ×<br>during | 0.714***<br>(0.171) | 0.224<br>(0.207) | 0.258<br>(0.173) | 0.002<br>(0.209) | 0.507**<br>(0.201) | 0.456**<br>(0.214) | 0.466**<br>(0.209) |
| Agreeableness<br>× during | −0.031<br>(0.101) | −0.128<br>(0.087) | −0.053<br>(0.094) | −0.055<br>(0.105) | −0.204**<br>(0.086) | −0.112<br>(0.088) | −0.098<br>(0.094) |
| Conscientious<br>× during | −0.118<br>(0.109) | −0.103<br>(0.096) | −0.091<br>(0.096) | −0.136<br>(0.098) | −0.146*<br>(0.080) | −0.205***<br>(0.077) | −0.064<br>(0.089) |
| Extraversion<br>× during | 0.085<br>(0.093) | 0.143<br>(0.093) | 0.145<br>(0.090) | 0.034<br>(0.089) | 0.146*<br>(0.075) | −0.017<br>(0.088) | −0.060<br>(0.089) |
| Neuroticism<br>× during | 0.013<br>(0.087) | 0.000<br>(0.082) | 0.048<br>(0.081) | −0.025<br>(0.084) | −0.045<br>(0.076) | −0.091<br>(0.082) | −0.061<br>(0.085) |
| Openness ×<br>during | 0.064<br>(0.089) | 0.215**<br>(0.095) | 0.172*<br>(0.089) | 0.201*<br>(0.105) | 0.209**<br>(0.098) | 0.271***<br>(0.099) | 0.309***<br>(0.102) |
| Cognitive Skills<br>× during | 0.065<br>(0.084) | 0.005<br>(0.129) | 0.129<br>(0.110) | 0.009<br>(0.121) | −0.063<br>(0.097) | 0.116<br>(0.113) | 0.088<br>(0.115) |
| Optimism ×<br>during | 0.017<br>(0.100) | 0.016<br>(0.093) | 0.007<br>(0.100) | −0.005<br>(0.100) | −0.003<br>(0.095) | −0.026<br>(0.102) | −0.095<br>(0.101) |
| Risk attitude<br>× during | −0.026<br>(0.036) | −0.033<br>(0.038) | −0.025<br>(0.038) | −0.048<br>(0.036) | −0.054<br>(0.035) | −0.028<br>(0.033) | −0.045<br>(0.036) |
| Locus of control<br>× during | 0.140**<br>(0.065) | 0.035<br>(0.069) | 0.038<br>(0.068) | 0.023<br>(0.075) | −0.016<br>(0.059) | −0.015<br>(0.064) | −0.020<br>(0.063) |
| *N* | 6,754 | 6,754 | 6,754 | 6,754 | 6,754 | 6,754 | 6,754 |

*Notes* Personality and Cognitive skills variables are standardized. All models control for individual fixed effects, job status, household income, any long term condition, month of the interview, age, region, marital status, household size and presence of children. Clustered standard errors at the individual level are in parentheses. \*\*\*, \*\*, and \* denote statistical significance at 0.01, 0.05, and 0.1 levels respectively.

**Table S.9:** Personality and Mental Health Deterioration During the Covid-19 Period, without sampling weights

|  | Dep. var. = GHQ12 (0 to 12) |  |  |  |  |  |  |
| --- | --- | --- | --- | --- | --- | --- | --- |
|  | (1) | (2) | (3) | (4) | (5) | (6) | (7) |
|  | 2019 and<br>Apr 2020 | 2019 and<br>May 2020 | 2019 and<br>Jun 2020 | 2019 and<br>Jul 2020 | 2019 and<br>Sep 2020 | 2019 and<br>Nov 2020 | 2019 and<br>Jan 2021 |
| post-covid | 0.952***<br>(0.215) | 0.432**<br>(0.214) | 0.194<br>(0.215) | -0.246<br>(0.211) | -0.370*<br>(0.217) | -0.078<br>(0.226) | 0.205<br>(0.231) |
| Female ×<br>post-covid | 0.685***<br>(0.088) | 0.515***<br>(0.088) | 0.397***<br>(0.091) | 0.215**<br>(0.087) | 0.393***<br>(0.088) | 0.468***<br>(0.092) | 0.511***<br>(0.092) |
| Agreeableness<br>× post-covid | -0.028<br>(0.048) | -0.083*<br>(0.048) | -0.075<br>(0.049) | -0.014<br>(0.046) | -0.008<br>(0.047) | -0.079<br>(0.049) | -0.102**<br>(0.049) |
| Conscientious<br>× post-covid | 0.004<br>(0.048) | -0.009<br>(0.048) | -0.034<br>(0.049) | -0.023<br>(0.047) | -0.093*<br>(0.048) | -0.079<br>(0.048) | -0.024<br>(0.049) |
| Extraversion<br>× post-covid | 0.045<br>(0.047) | 0.085*<br>(0.047) | 0.041<br>(0.047) | -0.008<br>(0.045) | 0.034<br>(0.045) | 0.019<br>(0.048) | 0.007<br>(0.047) |
| Neuroticism<br>× post-covid | -0.006<br>(0.047) | 0.006<br>(0.048) | 0.076<br>(0.049) | -0.029<br>(0.046) | -0.014<br>(0.048) | 0.000<br>(0.049) | 0.056<br>(0.050) |
| Openness ×<br>post-covid | 0.037<br>(0.046) | 0.095**<br>(0.047) | 0.157***<br>(0.049) | 0.106**<br>(0.047) | 0.132***<br>(0.047) | 0.150***<br>(0.049) | 0.147***<br>(0.049) |
| Cognitive Skills<br>× post-covid | 0.096**<br>(0.043) | 0.109**<br>(0.043) | 0.115***<br>(0.044) | 0.117***<br>(0.041) | 0.095**<br>(0.042) | 0.117***<br>(0.045) | 0.073*<br>(0.044) |
| *N* | 12,292 | 12,292 | 12,292 | 12,292 | 12,292 | 12,292 | 12,292 |

*Notes* Personality and Cognitive skills variables are standardized. All models control for individual fixed effects, job status, household income, any long term condition, month of the interview, age, region, marital status, household size and presence of children. Clustered standard errors at the individual level are in parentheses. \*\*\*, \*\*, and \* denote statistical significance at 0.01, 0.05, and 0.1 levels respectively.

**Table S.10:**
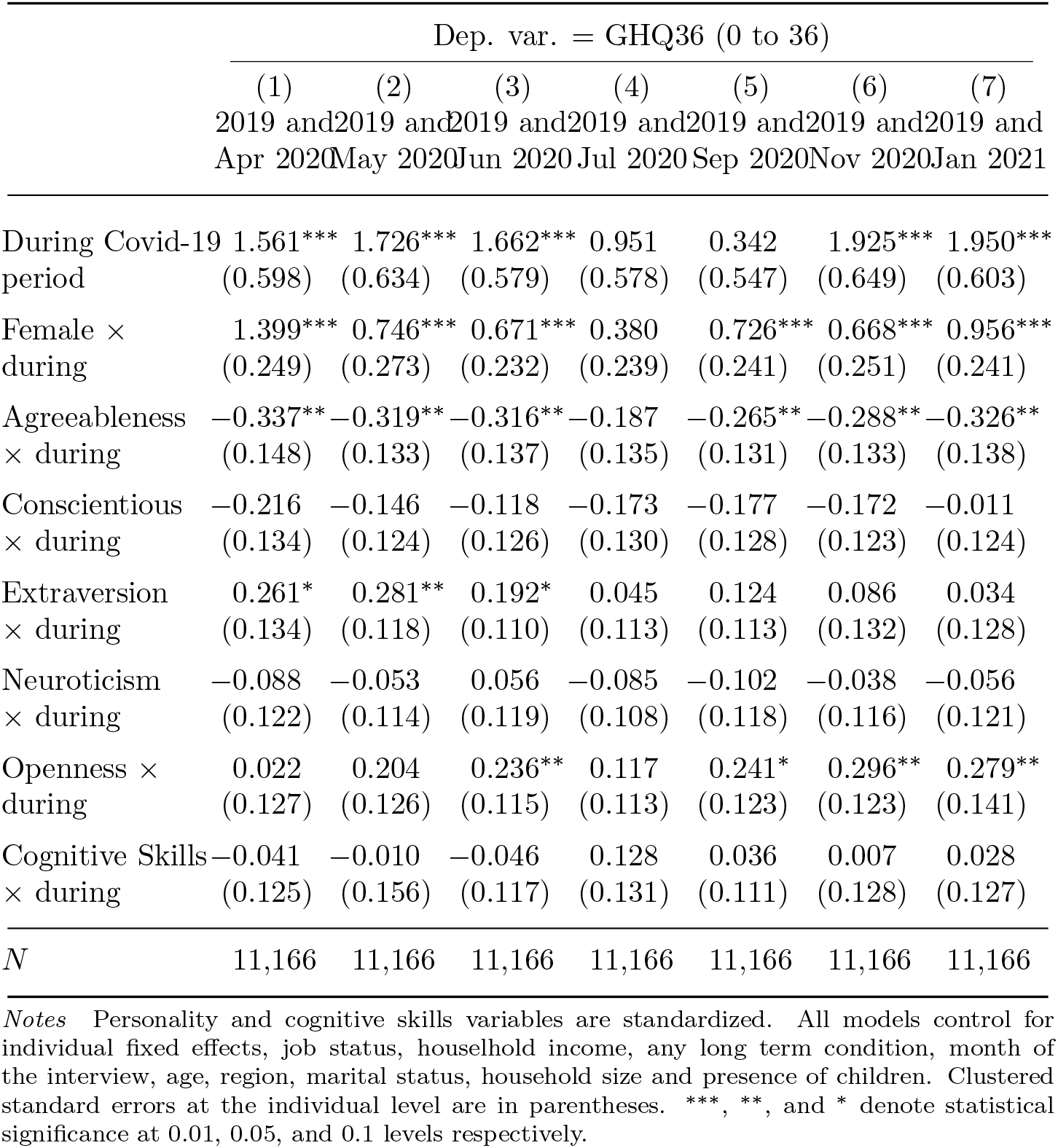
Personality and Mental Health Deterioration During the Covid-19 Period

**Table S.11:**
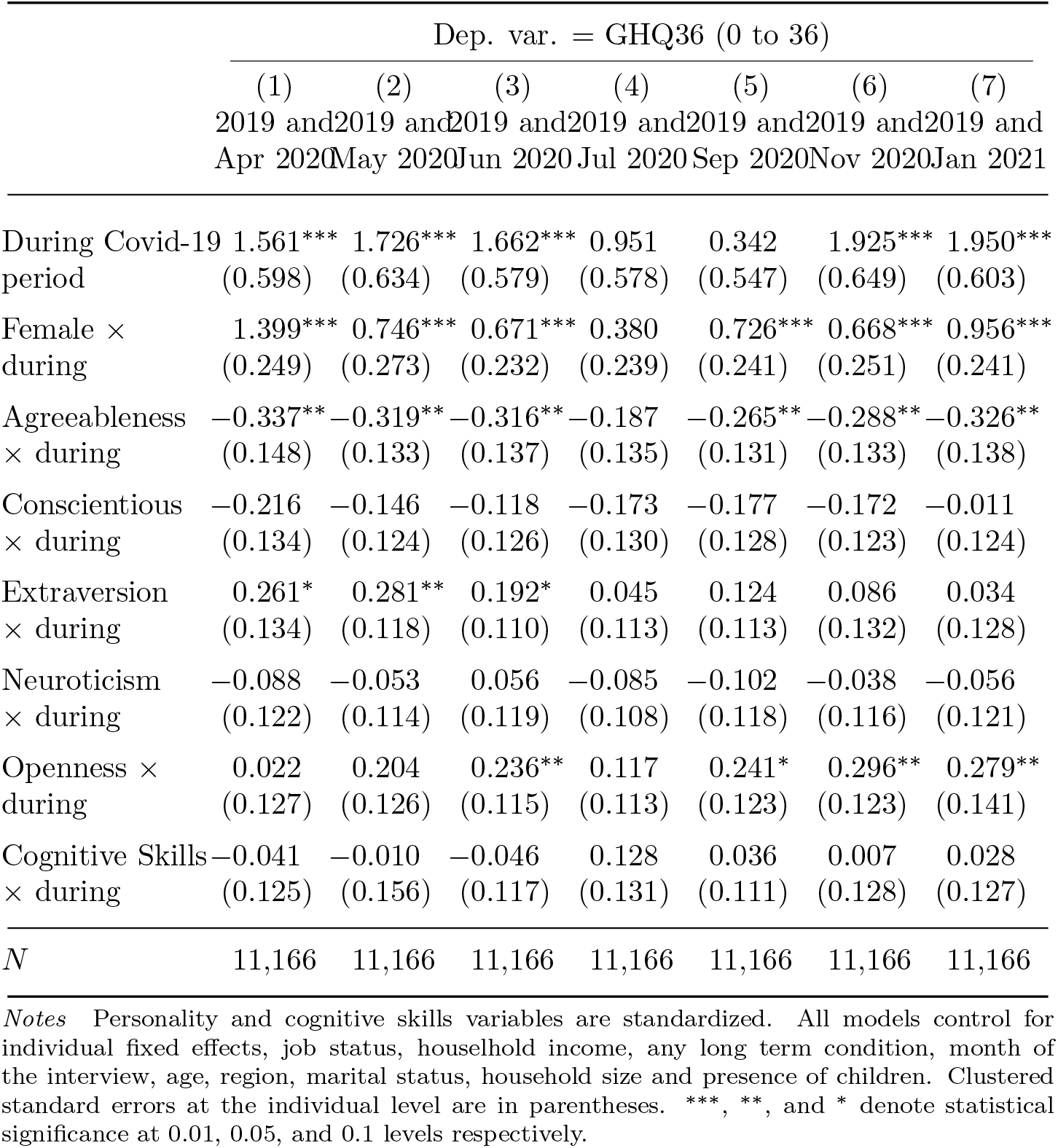
Personality and mental hHealth deterioration during the Covid-19 period, excluding over 60s and under 27s

**Table S.12:** Specification checks: personality and mental health deterioration during the Covid-19 period, June 2020

|  | Dep. var. = GHQ12 (0 to 12) |  |  |
| --- | --- | --- | --- |
|  | (1) | (2) | (3) |
|  | 2019 and<br>Jun 2020 | 2019 and<br>Jun 2020 | 2019 and<br>Jun 2020 |
| During Covid-19<br>period | 0.511***<br>(0.116) | 0.513***<br>(0.115) | 1.022***<br>(0.358) |
| Female ×<br>during | 0.514***<br>(0.151) | 0.527***<br>(0.155) | 0.513***<br>(0.154) |
| Agreeableness<br>× during | −0.216***<br>(0.072) | −0.210***<br>(0.073) | −0.192***<br>(0.070) |
| Conscientious<br>× during | −0.099<br>(0.079) | −0.097<br>(0.079) | −0.083<br>(0.076) |
| Extraversion<br>× during | 0.140*<br>(0.076) | 0.144*<br>(0.075) | 0.133*<br>(0.073) |
| Neuroticism<br>× during | −0.032<br>(0.067) | −0.028<br>(0.067) | −0.045<br>(0.066) |
| Openness ×<br>during | 0.148*<br>(0.076) | 0.130*<br>(0.075) | 0.145**<br>(0.073) |
| Cognitive Skills<br>× during |  | 0.082<br>(0.092) | 0.057<br>(0.088) |
| Controls | No | No | Yes |
| *N* | 11,166 | 11,166 | 11,166 |

*Notes* Personality and cognitive skills variables are standardized. Column (3) controls for individual fixed effects, job status, household income, any long term condition, month of the interview, age, region, marital status, household size and presence of children. Clustered standard errors at the individual level are in parentheses. \*\*\*, \*\*, and \* denote statistical significance at 0.01, 0.05, and 0.1 levels respectively.

**Table S.13:** Specification checks: personality and mental health deterioration during the Covid-19 period, January 2021

|  | Dep. var. = GHQ12 (0 to 12) |  |  |
| --- | --- | --- | --- |
|  | (1) | (2) | (3) |
|  | 2019 and<br>Jan 2021 | 2019 and<br>Jan 2021 | 2019 and<br>Jan 2021 |
| During Covid-19<br>period | 0.540***<br>(0.119) | 0.541***<br>(0.118) | 0.458<br>(0.365) |
| Female ×<br>during | 0.592***<br>(0.155) | 0.597***<br>(0.157) | 0.592***<br>(0.155) |
| Agreeableness<br>× during | -0.157**<br>(0.073) | -0.155**<br>(0.074) | -0.149**<br>(0.074) |
| Conscientious<br>× during | -0.005<br>(0.074) | -0.005<br>(0.074) | -0.013<br>(0.071) |
| Extraversion<br>× during | -0.059<br>(0.073) | -0.058<br>(0.072) | -0.054<br>(0.072) |
| Neuroticism<br>× during | -0.018<br>(0.070) | -0.017<br>(0.070) | -0.017<br>(0.069) |
| Openness ×<br>during | 0.220***<br>(0.073) | 0.213***<br>(0.077) | 0.228***<br>(0.076) |
| Cognitive Skills<br>× during | | 0.031<br>(0.086) | 0.044<br>(0.083) |
| Controls | No | No | Yes |
| *N* | 11,166 | 11,166 | 11,166 |

*Notes* Personality and cognitive skills variables are standardized. Column (3) controls for individual fixed effects, job status, household income, any long term condition, month of the interview, age, region, marital status, household size and presence of children. Clustered standard errors at the individual level are in parentheses. \*\*\*, \*\*, and \* denote statistical significance at 0.01, 0.05, and 0.1 levels respectively.

**Table S.14:** Mental health deterioration during the Covid-19 period among the top bottom 25% in Neuroticism

|  | Dep. var. = GHQ12 (0 to 12) |  |  |  |  |  |
| --- | --- | --- | --- | --- | --- | --- |
|  | (1) | (2) | (3) | (4) | (5) | (6) |
|  | 2019 and<br>Jun 2020 | 2019 and<br>Jun 2020 | 2019 and<br>Jun 2020 | 2019 and<br>Jan 2021 | 2019 and<br>Jan 2021 | 2019 and<br>Jan 2021 |
| During Covid-19<br>period |  | 0.557***<br>(0.090) | 0.364***<br>(0.121) |  | 0.755***<br>(0.102) | 0.455***<br>(0.134) |
| Female ×<br>during |  |  | 0.493***<br>(0.179) |  |  | 0.765***<br>(0.197) |
| Top 25% Neuroticism<br>× during | 0.829***<br>(0.152) | 0.272<br>(0.178) | 0.158<br>(0.173) | 1.011***<br>(0.173) | 0.257<br>(0.200) | 0.080<br>(0.193) |
| Cognitive Skills<br>× during | 0.084<br>(0.078) | 0.085<br>(0.077) | 0.110<br>(0.079) | 0.059<br>(0.089) | 0.061<br>(0.087) | 0.100<br>(0.086) |
| *N* | 8,365 | 8,365 | 8,365 | 8,365 | 8,365 | 8,365 |

*Notes* Only respondents scoring top and bottom 25% in Neuroticism are included. All models control for individual fixed effects, job status, household income, any long term condition, month of the interview, age, region, marital status, household size and presence of children. Clustered standard errors at the individual level are in parentheses. \*\*\*, \*\*, and \* denote statistical significance at 0.01, 0.05, and 0.1 levels respectively.

**Table S.15:**
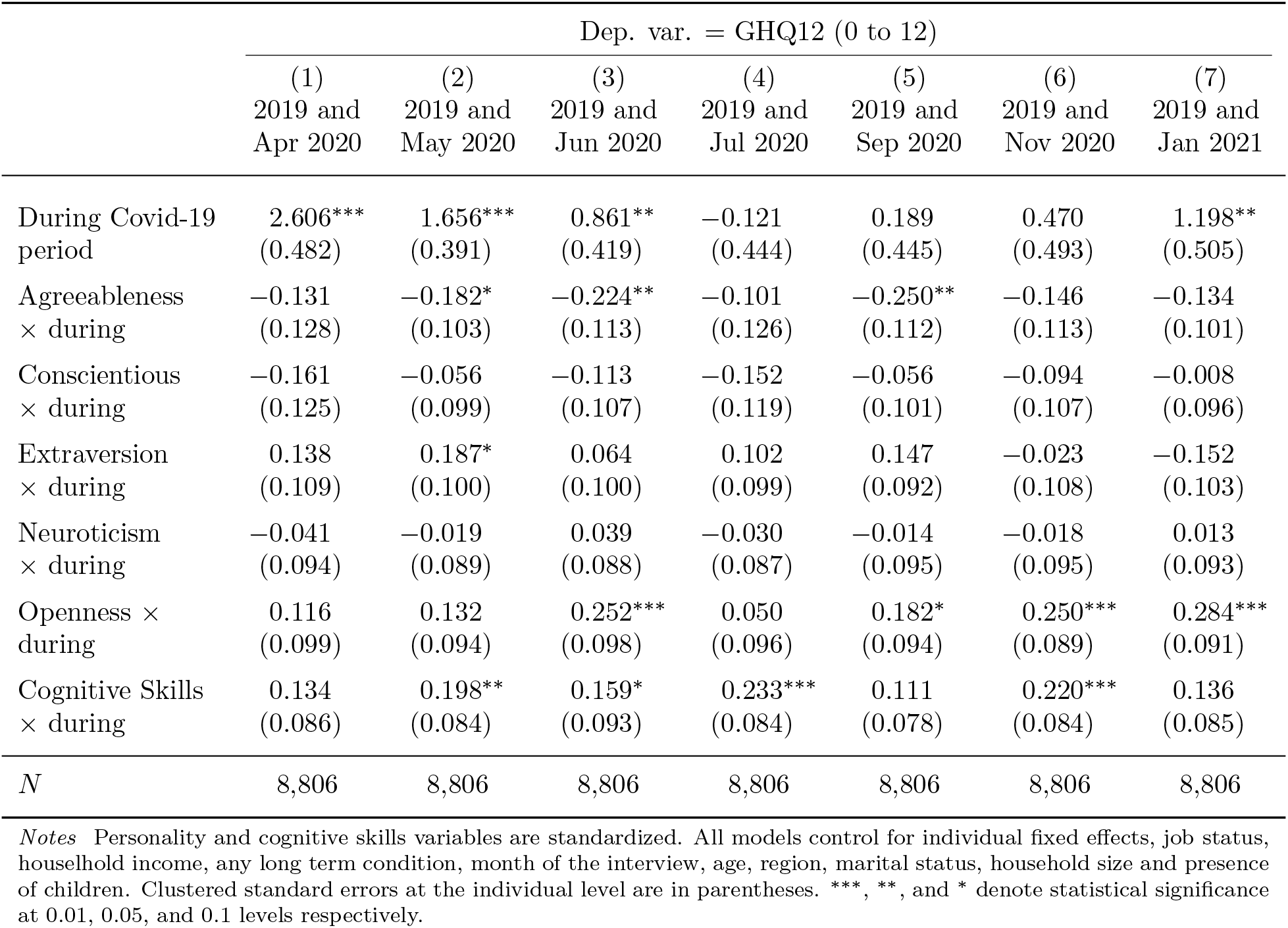
Personality and mental health deterioration during the Covid-19 Period, females only

**Table S.16:**
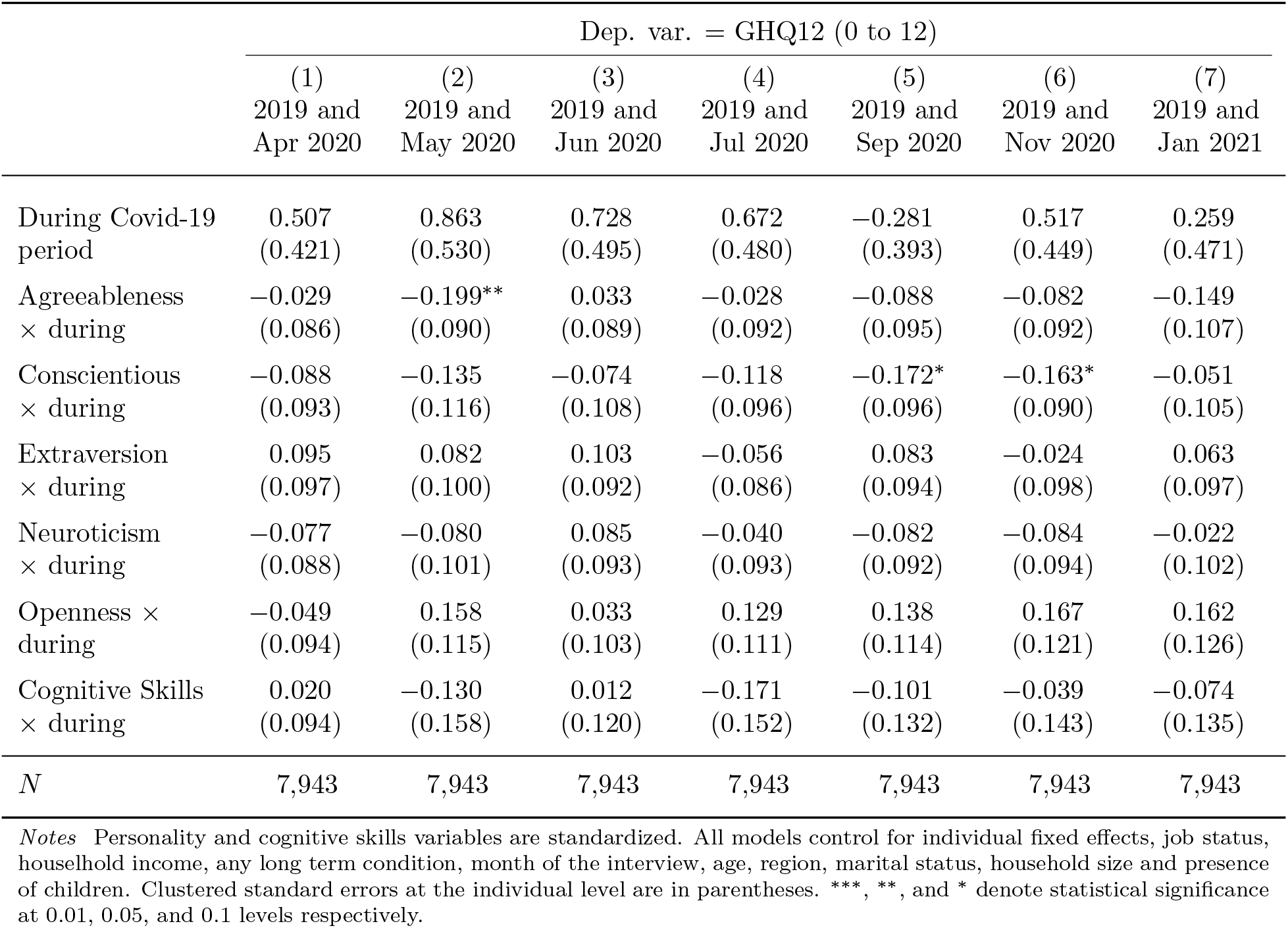
Personality and mental health deterioration during the Covid-19 period, males only

## Footnotes

1 A comprehensive review of this large literature is beyond the scope of this paper. We refer the reader to Kotov et al. (2010) for exhaustive meta-analysis and review of this literature, and to (Klein et al., 2011) for an illustration of the models linking personality to depression.

2 Furthermore, Cobb-Clark and Schurer (2012) show that they change very little even after very serious shocks like bereavement or unemployment

3 Men and women experience different kind of mental health problems. Men exhibit more externalizing disorders such as substance abuse and antisocial behavior (see e.g Rosenfield and Mouzon, 2013)

4 In SI Appendix, Table S4, we present similar estimations for each wave we use in this study; and in SI Appendix, Table S5, we run the same regressions by excluding all respondents older than 60 and younger than 27.

5 To understand better this apparent discrepancy with Figure 1, where there seems to be significant differences between top and bottom 25% Neuroticism scorers, in Appendix Table S15, we show that this difference vanishes once a general dummy variable indicating the Covid-19 period is introduced, suggesting that this effect is rather weak.

6 For expositional simplicity, We only included wave 2, 4 and 6 of the Covid-19 period, see SI Appendix Tables 15 and 16, for all waves.

